# Interferon-α2b treatment for COVID-19

**DOI:** 10.1101/2020.04.06.20042580

**Authors:** Qiong Zhou, Virginia Chen, Casey P. Shannon, Xiao-Shan Wei, Xuan Xiang, Xu Wang, Zi-Hao Wang, Scott J. Tebbutt, Tobias R. Kollmann, Eleanor N. Fish

**Author notes:** Address correspondence to: Eleanor N. Fish, Toronto General Hospital Research Institute, 67, College Street, Rm.4-424, Toronto ON M5G 2M1, Canada.

## Abstract

**Background:** The global pandemic of COVID-19 cases caused by infection with SARS-CoV-2 is ongoing, with no approved antiviral intervention. We describe here the effects of treatment with interferon-α2b in a cohort of confirmed COVID-19 cases in Wuhan, China.

**Methods:** In this retrospective study, 77 adults hospitalized with confirmed COVID-19 were treated with either nebulized IFN-α2b (5mU b.i.d.), arbidol (200mg t.i.d.) or a combination of IFN-α2b plus arbidol. Serial SARS-CoV-2 testing along with hematological measurements, including cell counts and blood biochemistry, serum cytokine levels, temperature and blood oxygen saturation levels were recorded for each patient during their hospital stay.

**Results:** Treatment with IFN-α2b with or without arbidol significantly reduced the duration of detectable virus in the upper respiratory tract and in parallel reduced duration of elevated blood levels for the inflammatory markers IL-6 and CRP.

**Conclusion:** These findings suggest that IFN-α2b should be further investigated as a therapy in COVID-19 cases.

## Introduction

In December 2019, an outbreak of pneumonia was reported in Wuhan, Hubei province, China, resulting from infection with a novel coronavirus (CoV), severe acute respiratory syndrome (SARS)-CoV-2. SARS-CoV-2 is a novel, enveloped betacoronavirus with phylogenetic similarity to SARS-CoV.^1^ Unlike the coronaviruses HCoV-229E, HCoV-OC43, HCoV-NL63 and HCoV-HKU, that are pathogenic in humans and are associated with mild clinical symptoms, SARS-CoV-2 resembles both SARS-CoV and Middle East respiratory syndrome (MERS), with the potential to cause more severe disease. A critical distinction is that CoVs that infect the upper respiratory tract tend to cause a mild disease, whereas CoVs that infect both upper and lower respiratory tracts (such as SARS-CoV-2 appears to be) may cause more severe disease. Coronavirus disease (COVID)-19, the disease caused by SARS-CoV-2, has since spread around the globe as a pandemic.

In the absence of a SARS-CoV-2-specific vaccine or an approved antiviral, a number of antivirals are currently being evaluated for their therapeutic effectiveness. Type I IFNs-α*/*β are broad spectrum antivirals, exhibiting both direct inhibitory effects on viral replication and supporting an immune response to clear virus infection.^2^ During the 2003 SARS-CoV outbreak in Toronto, Canada, treatment of hospitalized SARS patients with an IFN-α, resulted in accelerated resolution of lung abnormalities.^3^ ARB (Umifenovir) (ethyl-6-bromo-4-[(dimethylamino)methyl]-5-hydroxy-1-methyl-2 [(phenylthio)methyl]- indole-3-carboxylate hydrochloride monohydrate), a broad spectrum direct-acting antiviral, induces IFN production and phagocyte activation. ARB displays antiviral activity against respiratory viruses, including coronaviruses.^4^

Herein we report on the clinical course of disease in 77 confirmed cases of COVID-19 admitted to Union Hospital, Tongii Medical College, Wuhan, China, treated with interferon (IFN)-α2b, ARB, or a combination of IFN-α2b plus ARB.

## Materials & Methods

### Patients and treatments

Individuals with suspected COVID-19 were admitted to Union Hospital, Tongii Medical College, Wuhan, China, during the period January 16 - February 20, 2020, based on initial symptoms that included fever, chills, cough, sore throat, headache, nasal discharge, myalgia, fatigue, shortness of breath and/or diarrhea. We retrospectively examined the outcomes of patients with laboratory confirmed COVID-19 who received antiviral treatment with either IFN-α2b (Tianjin Sinobloway Biology, 5mIU/ml), arbidol (ARB) (arbidol hydrochloride; Jiangsu Simcere Pharm. Co.,100mg dispersible tablets), or a combination of IFN-α2b plus ARB, at the discretion of the treating physician, in accordance with the current practice guidelines. 5mIU IFN-α2b (1ml) were added to 2ml of sterile water and introduced as an aerosol by use of a nebulizer and mask. IFN-α2b treatment was bid, i.e. 10mIU/day. ARB treatment was 200mg (2 tablets) tid, i.e. 600mg/day. Additional COVID-19 confirmed cases from Wuhan Temporary Shelter Hospital (February 2-17, 2020), who were transferred to Union Hospital and treated with only ARB, were also included in this retrospective study. Ethics approval for analysis of all data collected was waived by hospital Institutional Review Boards, since all patient data collected conformed with the policies for a public health outbreak investigation of emerging infectious diseases issued by the National Health Commission of the People’s Republic of China.

### Laboratory tests

Throat swab specimens were tested by real time polymerase chain reaction (RT-PCR) for SARS-CoV-2. The SARS-CoV-2 laboratory test assay employed was based on the Centers for Diseases Control & Prevention, U.S.A. (CDC) recommendation.^5^ Briefly, throat-swab specimens from the upper respiratory tract of patients suspected of having SARS-CoV-2 infection were placed into collection tubes prefilled with 150µL of virus preservation solution and total RNA was extracted using a respiratory sample RNA isolation kit (High Pure Viral RNA Kit. Roche, Basel, Switzerland). RT-PCR assays for SARS-CoV-2 RNA were conducted using two target genes, namely open reading frame1ab (ORF1ab) and nucleocapsid protein (N). Samples were designated positive (+) or negative (-) based on a threshold adjusted to fall within the PCR exponential phase, for both target genes.^9^ Complete blood count and serum biochemical tests were assessed as per the Union Hospital’s routine clinical laboratory procedures. Serum cytokine levels (IL-2, IL-4, IL-6, IL-10, TNF-α, IFN-γ) were assayed using the BD Biosciences Th1/Th2 cytokine kit, as per the manufacturer’s instructions (BD Ltd., Franklin Lakes, NJ, USA) and peripheral blood cell populations enumerated using a BD FACSCanto Plus flow cytometer as per the Union Hospital’s routine clinical laboratory protocols.

### Statistical analysis

#### Time-to-event analysis

Time-to-viral clearance, defined as the number of days elapsed from the onset of symptoms to the time of the first of two consecutive negative PCR tests at least 24 hours apart, was compared between the treatment groups using time-to-event analysis. Time-to-viral clearance as a function of treatment was analyzed using Cox proportional hazards regression modelling, with age, coded as either a continuous variable or using categorical cut-offs of either >50 or >60 years of age, included as a covariate or not.

#### Time course analysis

Time course data were aligned to date of symptom onset and aggregated over 2-4-day intervals (depending on the analyte) to account for data not being available for all patients at all time points during disease course. For each interval and analyte, the statistical means were compared between treatment groups using analysis of variance (ANOVA). All analyses were carried out using R version 3.6.0.^6^

## Results

Table 1 summarizes the patient demographics and clinical characteristics of the cohort of COVID-19 cases evaluated in this retrospective study. 77 adults with confirmed COVID-19 admitted to Union Hospital, Wuhan, and at the discretion of the admitting physician, were treated with nebulized IFN-α2b (7), ARB (24) or a combination treatment of IFN-α2b plus ARB (46); IFN-α2b and ARB treatments were standard of care practice at this time in Wuhan. For 50% of all cases, treatment was started within 72hrs of confirmation of infection by a PCR(+) result; for 75% of cases, treatment started within 96hrs of a PCR(+) test and for 95% of cases, within 10 days of PCR(+). While all patients received various prophylactic antibiotic regimens as outlined in Supplementary Appendix, File 1, there was no case of proven or suspected bacterial infection.

**Table 1.** Demographics and clinical characteristics of patient cohort

|  | **IFN**<br>**n=7** | **IFN+ARB**<br>**n=46** | **ARB**<br>**n=24** | **P-value** |
| --- | --- | --- | --- | --- |
| Age, yrs | 41.3 (27-68) | 40.4 (25-80) | 64.5 (37-73) | <0.001 |
| Male (%) | 0 (0.0%) | 20 (43.5%) | 11 (45.8%) | 0.076 |
| Co-morbidities (%)<sup>1</sup> | 14.3% | 15.2% | 54.2% | 0.002 |
| **Symptoms on Admission:** |  |  |  |  |
| Fever (%) | 57.1% | 58.7% | 70.8% | 0.632 |
| Cough (%) | 42.9% | 50.0% | 54.2% | 0.888 |
| Fatigue (%) | 14.3% | 23.9% | 37.5% | 0.422 |
| Myalgia (%) | 14.3% | 13.0% | 29.2% | 0.228 |
| Headache (%) | 14.3% | 6.52% | 4.17% | 0.590 |
| Pharyngalgia (%) | 0.00% | 13.0% | 8.33% | 0.742 |
| Chest pain (%) | 14.3% | 6.52% | 20.8% | 0.134 |
| Expectoration (%) | 14.3% | 8.70% | 20.8% | 0.281 |
| Nausea (%) | 0.00% | 0.00% | 4.17% | 0.403 |
| Diarrhea (%) | 14.3% | 4.35% | 20.8% | 0.081 |
| Days from symptom onset to hospital admission<sup>2</sup> | 8.0<br>[5.5, 15.5] | 6.5<br>[3.0, 10.0] | 10.0<br>[4.5, 19.5] | 0.087 |
| Days from symptom onset to treatment<sup>2</sup> | 8.0<br>[6.5, 16.0] | 17.0<br>[10.0, 22.0] | 8.0<br>[5.0, 11.0] | 0.004 |

<sup>1</sup> Hypertension, diabetes, COPD, chronic bronchitis, heart disease, cancer<sup>2</sup> Median and interquartile range [Q1, Q3] is reported.

Serial clinical evaluations were performed on all patients (See Supplementary Appendix, File 1). Irrespective of the treatment group, none of the patients evaluated in this study exhibited persistent signs or symptoms of end organ dysfunction. Specifically, none of the patients developed respiratory distress requiring prolonged oxygen supplementation or intubation; consequently, none of the patients in this cohort required intensive care. Outside of the admission temperature, when approximately 50% of all patients exhibited fever (temperature > 38°C; which was successfully treated with ibuprofen), no other occurrence of fever was noted irrespective of antiviral treatment group (Supplementary Appendix, Figure 1). While all patients showed some radiographic abnormalities on chest computer tomography (CT) that were interpreted by local radiologists as ‘consistent with viral pneumonia,’ detailed evaluation of the CT findings were not performed due to the overwhelming workload at Union Hospital at the time of this study. Serial laboratory measurements of blood levels for haemoglobin, glucose, total bilirubin, direct bilirubin, alanine aminotransferase (ALT), aspartate aminotransferase (AST), lactate dehydrogenase (LDH), creatine kinas (CK), blood urea nitrogen (BUN), albumin (Alb), creatinine, and troponin 1 were also conducted (Supplementary Appendix, Figure 2). Beyond a mild transaminitis (ALT elevation) early during hospitalization, which subsequently improved in all patients, the data for blood chemistries indicated that levels fluctuated closely around the limits of normal over the course of hospitalization, without a clear or consistent difference among treatment groups. Peripheral blood cell populations, including total white blood cells (WBC), lymphocyte, CD4+ T cell, CD8+ T cell, B lymphocyte, neutrophil, NK cell and platelet counts were also measured over the course of hospitalization (Supplementary Appendix, Figure 3). With the exception of elevated platelets, which peaked two weeks into the disease course, all other cell populations fluctuated around the normal range with no clear or consistent difference discernible among antiviral treatment groups. Together, the clinical and laboratory data indicate that the entire cohort evaluated in this study consisted of moderate cases of COVID-19 across all treatment groups.

**Figure 1.**
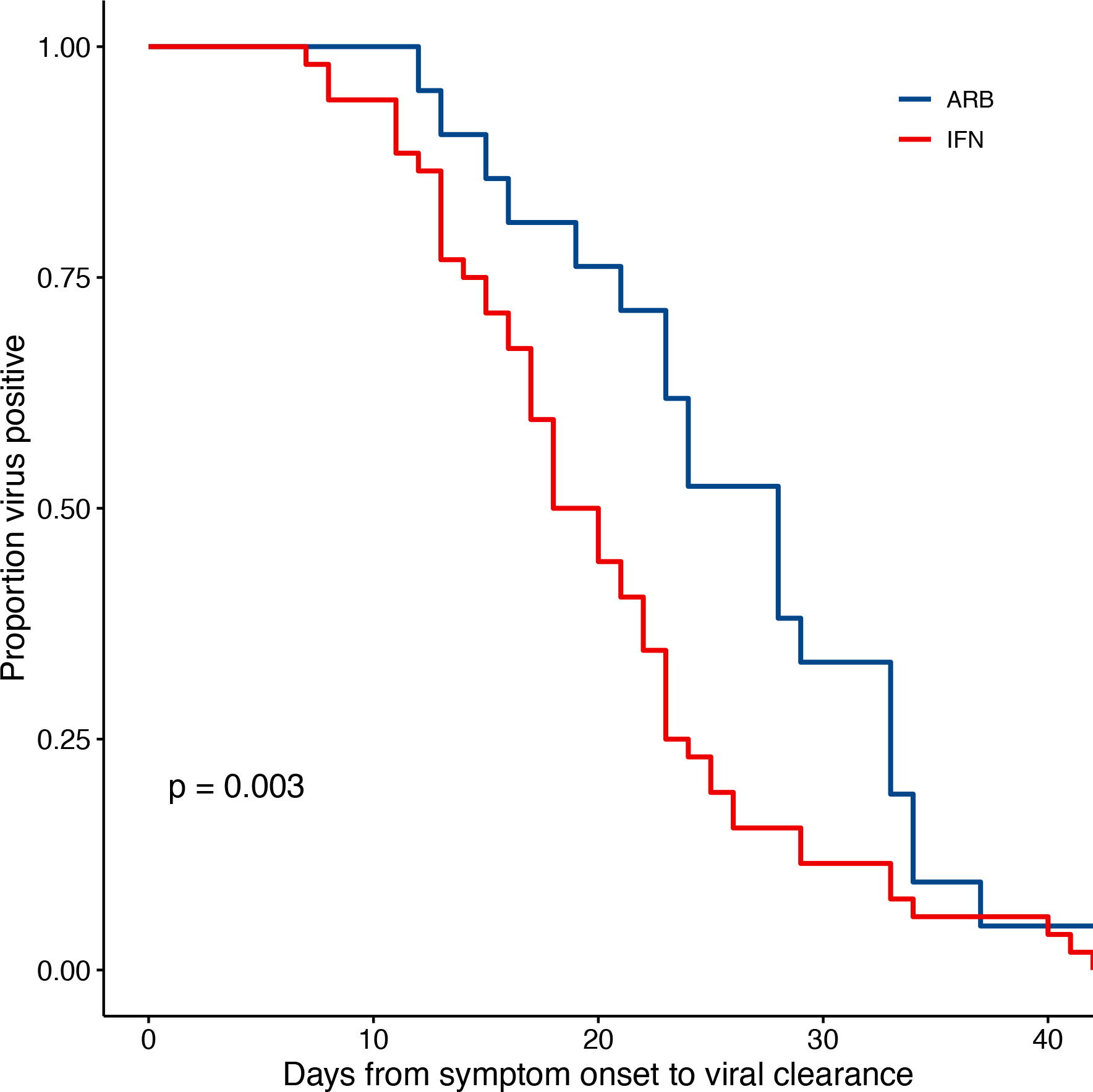
IFN-α2b treatment accelerated viral clearance. Confirmed COVID-19 cases were treated either with ARB alone (ARB; 24 patients) or IFN-α2b with or without ARB (IFN; 53 patients). Upper respiratory samples were assessed by PCR for the presence of SARS-CoV-2. Shown is the proportion of patients that had detectable virus as a function of the day of sampling from symptom onset.

**Figure 2.**
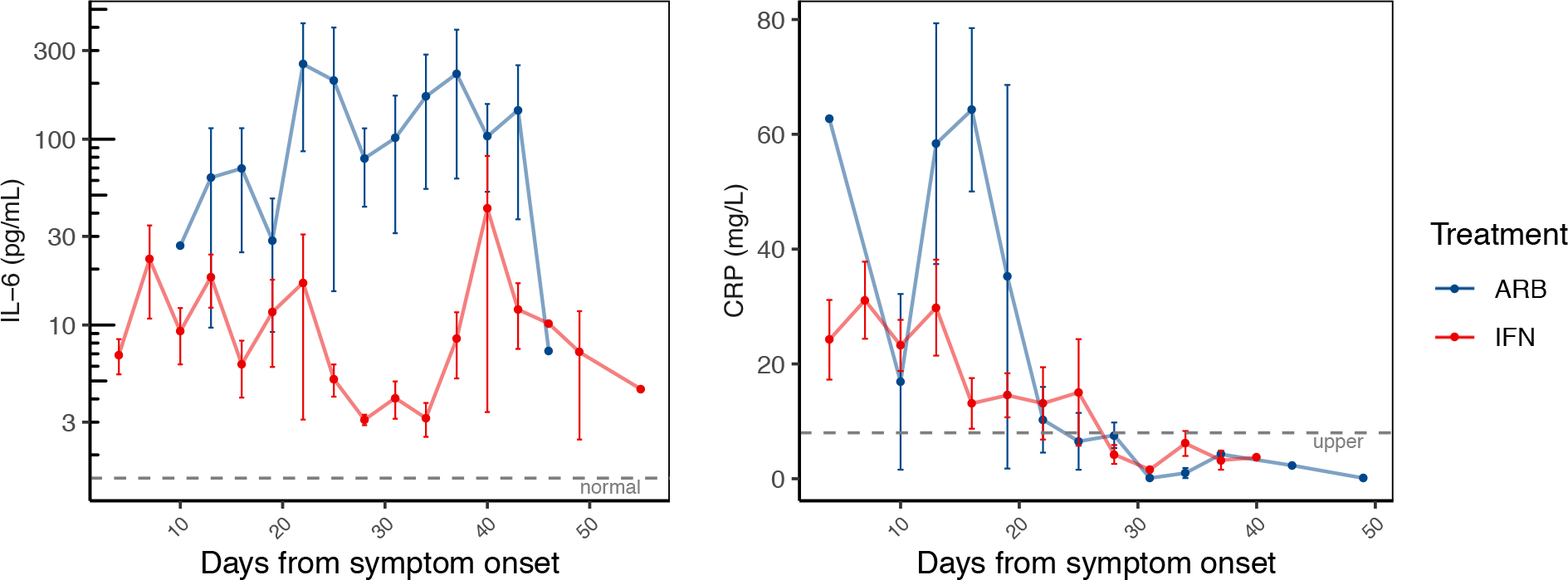
Reduced inflammatory markers with IFN-α2b treatment. The same patients as in Figure 1 were serially sampled for assessment of interleukin-6 (IL-6; panel **A)** and C-reactive protein (CRP; panel **B)** from the day of symptom onset. Values recorded were aggregated across 3 day intervals and shown as the mean +/- SE

Clinical course of the COVID-19 cases was also assessed in relation to age, sex and co-morbidities. With the exception of haemoglobin, which was lower in females, for each of the other measurements listed above we found no effect of age, sex or co-morbidities on disease course or laboratory measurements.

Viral clearance was defined as two consecutive negative PCR tests at least 24 hours apart as previously described.^5^ Assessing disease course from Day of symptom onset (D0) to the first negative (-) PCR of 2 consecutive PCR (-), the data revealed a significantly different rate of viral clearance for each treatment group (Supplementary Appendix, Figure 4). Specifically, outcome analysis suggested that treatment with IFN-α2b, whether alone or in combination with ARB, accelerated viral clearance when compared to ARB treatment alone. Closer scrutiny of the treatment regimens for those cases treated with a combination of IFN-α2b and ARB revealed that for 16 of the 46 cases (34.8%) IFN-α2b treatment was started after ARB treatment had been initiated and, for 24 cases (52.2%), IFN-α2b treatment was continued after ARB treatment was stopped (Supplementary Appendix, Figure 5). Given the heterogeneity of treatment regimens within this treatment group, we considered the time to viral clearance for all cases treated with IFN (i.e. combined the IFN-only with the IFN plus ARB groups) compared to those who received ARB only. The data shown in Figure 1display the statistically significant accelerated viral clearance from the upper respiratory tract in patients who received IFN-α2b treatment (p=0.003).

Circulating cytokine levels (IL-2, IL-4, IL-10, IFN-γ, IL-6, TNFα) and biomarkers of inflammation (C-reactive protein, CRP and procalcitonin, PCT) were also examined over the disease course. Circulating levels of PCT, IL-2, IL-4, IL-10, IFN-γ and TNFα remained within their normal range throughout disease course, irrespective of treatment group (Supplementary Appendix, Figure 6). Notable and significant exceptions were IL-6 and CRP. As disease course progressed and prior to resolution, we observed a clear distinction of serum IL-6 levels between cases treated with IFN (i.e. IFN alone or IFN & ARB) and cases treated with ARB alone. More specifically, whereas circulating levels of IL-6 remained low for all patients who received IFN, those who received ARB alone (i.e. with no IFN) exhibited a significant spike in circulating IL-6 levels (Figure 2A, Supplementary Appendix, Figure 7). We also noted elevated levels of CRP over approximately the same time that IL-6 was elevated (Supplementary Appendix, Figure 7). Similar to IL-6, CRP also returned to within normal range as disease resolved. These data suggested that treatment with IFN, whether alone or in combination with ARB, limited the circulating CRP level (Figure 2B).

Sex and co-morbidities had no effect on the effects of ARB and IFN treatment on time to viral clearance, or IL-6 and CRP levels. Cognizant that the ARB-only treatment group consisted generally of older patients, we adjusted for age in the statistical analyses. Regardless of whether age was considered as a continuous variable or a categorical variable (<50 yrs vs ≥50 yrs; <60 yrs vs ≥60 yrs), the effects of IFN treatment on IL-6, CRP, and time to viral clearance remained statistically significant. For those cases treated with ARB alone, IL-6 levels were significantly higher than for those treated with IFN from day 12-47 (p=2.2 × 10^−9^). Similarly, for the ARB alone treatment group, CRP levels were significantly higher than for those cases treated with IFN from day 0-20 (p=0.0033). Time to viral clearance was significantly shorter for those cases treated with IFN-α2b compared to those treated only with ARB (p=0.003).

## Discussion

This retrospective study provides several important and novel insights into COVID-19 disease. Most importantly, IFN-α2b therapy appears to shorten duration of viral shedding. Notably, reduction of markers of acute inflammation such as CRP and IL6 correlated with this shortened viral shedding, suggesting IFN-α2b acted along a functional cause-effect chain where virally induced inflammation represents a pathophysiological driver. Taken together, these findings elevate the biological plausibility of IFN-α2b representing a therapy for COVID-19 disease.

As the SARS-CoV-2 pandemic takes an ever-increasing toll, the urgent search for effective prophylactic and therapeutic interventions is rapidly accelerating. This includes lopinavir/ritonavir,^7,8^ chloroquine,^9^ remdesivir^,10^ as well as interferon (IFN)-α^2^ and arbidol (ARB)^4^ and combinations of these. Most of these antivirals only have *in vitro* data to support consideration for coronavirus targets prior to clinical testing; as such, while unfortunate, it is not surprising that there is a high chance of failure.^11^ However, we had shown during the SARS-CoV-1 outbreak in Canada that IFN-α treatment could hasten resolution of coronavirus-mediated human disease.^3^ This prompted us to evaluate IFN-α therapy that was administered for COVID-19 disease in the early stages of the outbreak in Wuhan, Hubei province, China. Indeed, our analysis suggests that inhaled IFN-α2b accelerated viral clearance from the respiratory tract and hastened resolution of systemic inflammatory processes when compared to ARB treatment alone. While we recognize that these data are at best suggestive, given the urgency, the findings indicate that a follow-up randomized placebo controlled clinical trial (RCT) is now warranted. Success could not only benefit the individual infected patient but, by reducing duration of viral shedding even in moderate cases (such as this cohort), assist in slowing the population spread.

The reduction of the inflammatory biomarker IL-6 following inhaled IFN-α2b therapy not only supported a clinically relevant impact of this approach, but also hinted at likely functional connections between viral infection and host end organ damage. IL-6 has been shown to provide prognostic value in acute respiratory distress syndrome (ARDS), which is the most severe form of COVID-19 disease.^12^ If this were indeed the case, then targeting interventions such as interleukin-6 (IL-6) receptor inhibitors (e.g. toclizumab or sarilumab) towards this axis may prove a useful therapeutic adjunct, at least in those most severely ill. This form of therapy has recently been approved by China’s National Health Commission^13^ and is currently under consideration by the Italian Medical Agency.^14^ The advantage of IFN-α2b over blocking IL-6 rests in IFN targeting the cause (SARS-CoV-2), not only the symptoms (IL-6).

This retrospective study has several significant limitations. Most obvious is the fact that the study cohort was small, non-randomized, with unbalanced demographics between treatment arms that were of unequal size. However, we considered this an exploratory study only, with the objective of determining in as rapid a manner as possible if a full trial should be considered. The results indicate that an IFN-α RCT is now warranted. Furthermore, since the entire cohort consisted only of moderate cases of COVID-19 disease, our findings may not be indicative of what occurs in more severely ill patients; such caution about generalizability is indeed further supported by the lack of impact of age, sex and comorbidities on the course of COVID-19 disease in our cohort, as all these have been shown to influence clinical course.^15^

Irrespective of these significant limitations, to our knowledge, the findings presented here are the first to suggest therapeutic efficacy in COVID-19 disease of IFN-α2b, an available antiviral intervention. Furthermore, beyond clinical benefit to the individual patient, treatment with IFN-α2b may also benefit public health measures aimed at slowing the tide of this pandemic, in that duration of viral shedding appears shortened.

## Data Availability

Data are available

## Contributors

QZ was responsible for patient care and treatment, clinical oversight and clinical data collection. VC and CPS analysed the data and generated the figures. X-SW, XX, XW and Z-HW collected laboratory and radiographic data. SJT analysed data. TRK, ENF conducted data analysis, data interpretation, literature searches and manuscript writing.

